# DDASR: Domain-Distance Adapted Super-Resolution Reconstruction of MR Brain Images

**DOI:** 10.1101/2023.06.29.23292026

**Authors:** Shan Cong, Kailong Cui, Yuzun Yang, Yang Zhou, Xinxin Wang, Haoran Luo, Yichi Zhang, Xiaohui Yao

## Abstract

High detail and fast magnetic resonance imaging (MRI) sequences are highly demanded in clinical settings, as inadequate imaging information can lead to diagnostic difficulties. MR image super-resolution (SR) is a promising way to address this issue, but its performance is limited due to the practical difficulty of acquiring paired low- and high-resolution (LR and HR) images. Most existing methods generate these pairs by down-sampling HR images, a process that often fails to capture complex degradations and domain-specific variations. In this study, we propose a domain-distance adapted SR framework (DDASR), which includes two stages: the domain-distance adapted down-sampling network (DSN) and the GAN-based super-resolution network (SRN). The DSN incorporates characteristics from unpaired LR images during down-sampling process, enabling the generation of domain-adapted LR images. Additionally, we present a novel GAN with enhanced attention U-Net and multi-layer perceptual loss. The proposed approach yields visually convincing textures and successfully restores outdated MRI data from the ADNI1 dataset, outperforming state-of-the-art SR approaches in both perceptual and quantitative evaluations. Code is available at https://github.com/Yaolab-fantastic/DDASR.

## I. INTRODUCTION

Magnetic resonance imaging (MRI) is widely used as a crucial neuroimaging method because it delivers detailed insights into brain tissue structure without the need for invasive procedures. However, obtaining high-quality MRI images often involves challenges such as the need for extended scanning times, high magnetic field strengths, and patient discomfort, which can lead to motion-sensitive images and increased costs [1]. Additionally, the complex nature of MR signals makes them prone to various distortions and artifacts, including motion blur, noise, and signal dropout. These further complicate the imaging process and potentially hinder accurate diagnoses. Single image super-resolution (SISR) techniques have been developed to enhance a low-resolution (LR) image by improving signal-to-noise ratio [2], accentuating texture details, and eliminating visual artifacts [3]. This technology is particularly beneficial in brain imaging as it offers a solution to the labor-intensive and demanding process of analyzing substandard MR images that lack sufficient information. While recent advances in deep learning-based MR image SR offer a promising solution to enhance image quality and maintain diagnostic quality [4], there are several challenges that must be addressed before clinical implementation.

One major challenge is modeling the unknown and complex degradation of a real LR image in the absence of paired LR/HR MR images. Since paired LR/HR MR images are usually unavailable, existing methods are dedicated to generating an LR/HR pair by synthesizing an LR image from the HR image to model the transformation [5], [6]. Recent works primarily focused on explicit modeling (as shown in Fig. 1(a)), such as bicubic down-sampling [7], k-space filtering [8], and combination of image degradation operations [5], [9]. These works performed well when the true degradation is known prior. However, the performance is significantly compromised when the real degradation differs from their estimation. Alternatively, a few other works employ implicit modeling (as shown in Fig. 1(b)), such as domain-distance adaptation using convolutional neural network (CNN) [6], data distribution learning with GAN [10], and representation learning [11]. *Although they offer greater flexibility in handling various degradation scenarios, as shown in Fig. 1(b), existing implicit methods primarily focus on minimizing the distance between real and synthetic images and overlook the transfer of domain representations*.

**Fig. 1.**
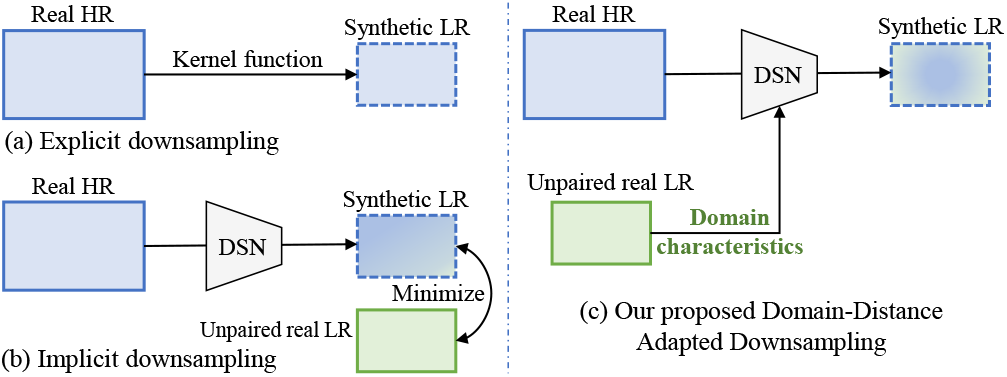
Synthesizing downsampled LR images from the real HR and unpaired real LR datasets. (a) shows the explicit down-sampling. (b) shows the conventional implicit down-sampling, which proposes minimizing the distance between synthetic and real LR images. (c) shows our proposed domain-distance adapted down-sampling method, in which real LR domain characteristics were embedded into the down-sampling process to facilitate the domain translation.

Another challenge is recovering texture details while preserving the topological structures. An enhanced image may pass quantitative assessment but fail visual inspection as existing algorithms usually fail in balancing sharpness and structural details [12]. In some cases, tissue structure is even altered, which can be a fatal error as the structural texture is critical for visual diagnostics (i.e., lesion localization) and neuroscience studies (i.e., brain parcellation). There are two common directions to prioritize the preservation of topological structures and texture details: 1) design proper deep learning architectures for SR, where recent efforts have been made to improve autoregressive models [13], GAN [14], [15], variational autoencoder (VAE) [16], [17], encoding-decoding networks [18], [11], and diffusion probabilistic models [19]; and 2) design loss functions that consider visual effects to produce convincing results like perceptual loss [14], [7] and texture matching loss [20], [21] functions. Overall, the investigation in both directions offers exciting opportunities for advancing MR image SR, and we believe that their combination has the potential to yield even more impressive results.

To tackle the aforementioned challenges, we develop a realistic down-sampling strategy, as illustrated in Fig. 1(c). This strategy aims to incorporate LR domain characteristics while learning the mapping relationship between the target domain (i.e., real LR) and the source domain (i.e., real HR). In addition, it includes an effective texture restoration component that utilizes multi-scale image features for enhancing MR brain image super-resolution. By combining these innovative designs, our proposed method significantly improves the preservation of both structural integrity and texture details, leading to better visual and quantitative quality. Our contributions can be summarized as follows:

- **Realistic down-sampling:** We introduce a transformer-based, domain-distance adapted down-sampling network (DSN) to efficiently capture domain representations and incorporate these features into the mapping relationship between HR images and LR images.
- **Structure and texture preservation:** We propose a super-resolution network (SRN) that employs a generator enhanced with multi-layer perceptual loss and a discriminator utilizing an attention U-Net to produce biologically reasonable textures and precise contours.
- **Denoising and super-resolution:** We propose a novel domain-distance adapted SR framework to enhance both the image quality and resolution of MR brain images, achieving substantial improvements over state-of-the-art models in both qualitative visual assessments and quantitative evaluations.

## II. Methodology

Given two MR image domains characterized by two sets of unpaired HR images 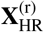 and LR images 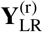, our goal is to train an SR model that can increase the resolution of an LR domain image while enabling the synthesized HR image align with the real HR domain. To achieve this, we propose a two-stage framework that comprises a down-sampling network and a super-resolution network.

The down-sampling stage (Fig. 2(a)) employs a transformer architecture to embed the real LR domain representation into the downsampled images and introduces the content-aware positional encoding (CAPE, Fig. 2(c)) to capture invariant brain structural representation from source HR images and preserve fine-grained details. The SR stage (Fig. 2(b)) uses a GAN architecture, where the generator (Fig. 2(d)) employs an encoder-bottleneck-decoder scheme and the discriminator (Fig. 2(e)) adopts an attention U-Net. Meanwhile, to ensure visually convincing textures, we incorporate multi-layer perceptual loss into the SR stage. We present the details of the domain-distance adapted down-sampling strategy in Sect. II-A, the GAN-based SR framework in Sect. II-B for reproducibility. In principle, one can apply any image SR task in the proposed architecture.

**Fig. 2.**
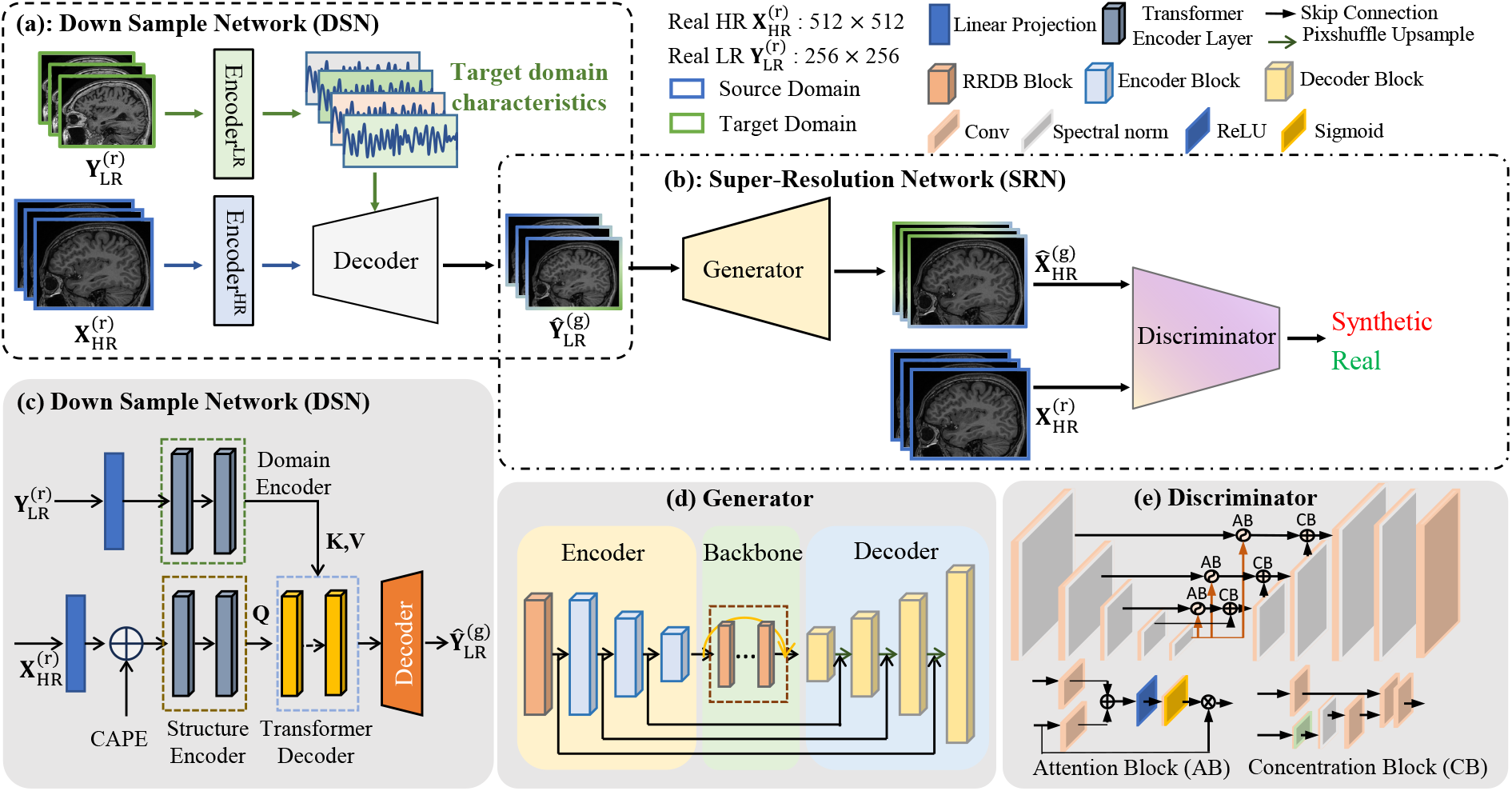
Overview of the proposed two-stage SR framework. (a) Stage 1: taking the real HR and real unpaired LR data as input, a transformer-based down-sampling network (DSN) is designed to generate paired (to the HR images) LR images, aligning to the real LR domain. (b) Stage 2: the synthesized (LR, HR) pairs are used to train the super-resolution network (SRN), where the GAN architecture with multi-layer perceptual loss (for the generator) and attention U-Net mechanisms (for the discriminator) is employed. (c-e) illustrate the details of DSN, GAN generator, and discriminator, respectively.

### A. Domain-Distance Adapted Down-Sampling

Existing down-sampling methods, such as linear, k-space, blur kernels, or a combination of down-sampling operations have low generalization ability and cannot adapt to complex degradation processes during MRI scans. To address this, we propose a domain distance-adaptive down-sampling network (DSN) based on the transformer architecture. As shown in Fig. 2(c), we employ transformer blocks with attention mechanisms to extract image domain characteristics, textures, and structural features, thereby facilitating the construction of source and target feature embeddings in the latent space [22]. This mechanism enables the transformer to capture local and global contextual information and generate multi-scale feature representation. In addition, we apply CAPE [23] on the source HR images 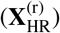 during multi-scale down-sampling, which aims to ensure that the brain structure and texture information remains invariant during the transformation process by encoding the positional information.

#### Image positional coding

Given a source image 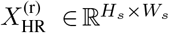 and a target image 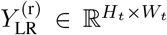, patches are projected into a sequential feature embedding *E* of size *N* × *C* using a linear projection layer, where 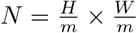 and *C* are the length and dimension of *E* respectively, and *m* × *m* is the patch size. The CAPE *P*_*c*_ was proposed to integrate the semantics of image content into the positional encoding, thereby reducing the impact of the image scale on the relative distance. Specifically, *P*_*c*_(*i, j*) between patches (*i, j*) is calculated as follows:

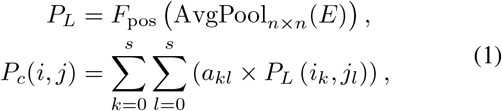

where *F*_pos_ is a 1 × 1 convolution operation, *P*_*L*_ is the learnable relative positional relationship, *n* is the block size, *a*_*kl*_ is the interpolation weight, and *s* is the number of adjacent patches.

#### Transformer encoder

Two transformer encoders are employed to encode the structure information from the source domain image 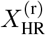 and encode the domain characteristic information from the target domain image 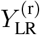. Specifically, the embeddings of the source domain image sequence *Z*_*s*_ = {*E*_*s*1_ + *P*_*c*1_, *E*_*s*2_ + *P*_*c*2_, · · ·, *E*_*sL*_ + *P*_*cL*_, } and the target domain image sequence *Z*_*t*_ = {*E*_*t*1_, *E*_*t*2_, · · ·, *E*_*tL*_, } are fed into each transformer encoder to produce the encoded struc-ture sequence 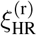 and domain characteristic sequence 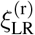.

Each encoder layer is composed of a multi-head attention (MHA) module and a feed-forward network (FFN) [24], which together encode the input sequence as follows:

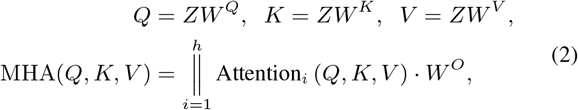

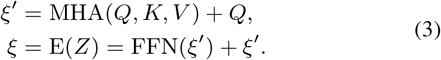

For each input sequence, the queries (Q), keys (K), and values (V) are generated via three learnable weight matrices *W* ^*Q*^, *W* ^*K*^, and *W* ^*V*^, respectively. The ∥ denotes the concatenation of *h* independent heads, and *W* ^*O*^ is the projection matrix that aggregates the output from all attention heads.

Following Eq. 2 and Eq. 3, the source HR images and the target LR images are encoded as 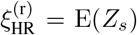 and 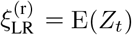.

#### Transformer decoder

As shown in Fig. 2(c), the transformer decoder is used to generate decoded image 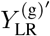 from the source image sequence 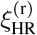 while absorbing the domain characteristic information from the target image sequence 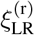. In particular, the transformer decoder consists of two cross-attention layers and one FFN, from which the decoded image 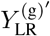 is calculated as:

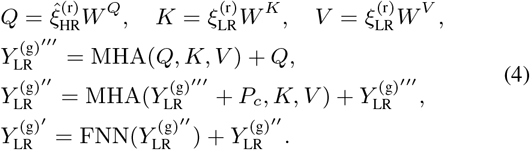

Here, 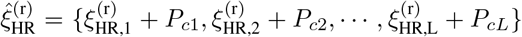 is the CAPE encoded source image sequence.

#### Resizing

For the training purposes, the decoded output 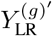 undergoes additional processing through a three-layers up-sampling decoder [25] and a linear degradation operation to obtain the downsampled result 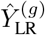. There is a convolution layer of 3 × 3 convolution kernel, ReLU activation function, and 2× upsample applied in each three-layer convolution up-sampling module, respectively. By controlling the number of layers of the CNN up-sampling module, we can output images with any integer multiple downsampled multiple.

#### DSN loss

There are two objectives for down-sampling: preserving the original structure and texture of the source HR MR image and attaining the domain characteristics of the target LR images. To achieve these, two loss functions are introduced: the structural loss ℒ_*s*_ that reduces the structure and texture discrepancies between the generated LR 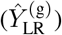 and real HR images 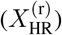, and the domain loss ℒ_*d*_ to address the gap between synthesized LR and target LR domains (i.e., real LR domain). To improve the model stability, we employ the pre-trained VGG19 to extract feature maps and train the model:

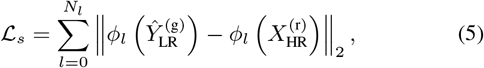

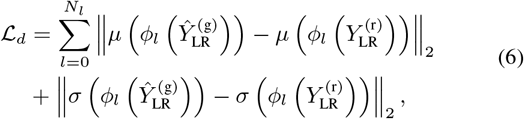

where *N*_*l*_ denotes the number of VGG layers, and *ϕ*_*l*_(·) extracts features from the *l*-th VGG layer, *µ*(·) and *σ*(·) represent the mean and variance of the features, respectively.

To help keep more detailed and local structural information, we further introduce an identity loss ℒ_*id*_ into the DSN optimization. Instead of trading off between image structure and domain, the identity loss considers both local mapping and global distributions by maintaining structural information rather than changing domain representation. Let 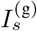 and 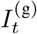 denote the images generated by inputting two identical source domain images *I*_*s*_ and two identical target domain images *I*_*t*_ into the DSN, respectively. The identity loss measures the difference between *I*_*s*_ and 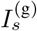 for the source domain, and between *I*_*t*_ and 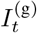 for the target domain:

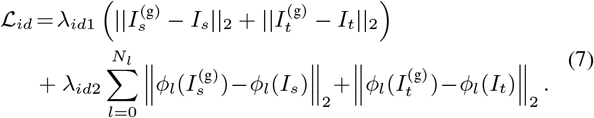

It should be noted that all images are resized to have the same dimension during the training process.

Overall, the total DSN loss can be expressed as:

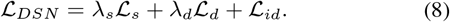

Empirically, we set *λ*_*s*_ = 7, *λ*_*d*_ = 10, *λ*_*id*1_ = 30 and *λ*_*id*2_ = 5 across the experiments.

### B. GAN-based super-resolution

Using the pseudo (LR, HR) image pairs synthesized by the DSN, a GAN-based super-resolution network (SRN) is trained to model the HR reconstruction. We now detail the SRN framework, as illustrated in Fig. 2(b,d,e).

#### Generator

The generator consists of an encoder, a bottleneck, and a decoder (see Fig. 2(d)). The encoder extracts features from the input LR images, the bottleneck incorporates dense connections and deeper networks for deep feature fusion, and the decoder merges features from both the encoder and bottleneck to generate HR images. This process emphasizes the restoration of both large-scale contour structures and fine-grained texture details.

##### Encoder

We employ the residual-in-residual dense block (RRDB) [14] for effective feature extraction, which first extracts features from the LR image, yielding a low-dimensional feature representation *f*_0_. To capture multi-scale feature information, we then employ convolutional modules that progressively downsample the features while expanding their dimensionality. This process yields multi-scale features *f*_*i*_ = *E*_*i*_ (*f*_*i*−1_), *i* ∈ {1, · · ·, *N*}, where *E*_*i*_ (·) represents a convolutional layer with a kernel size of 4, a stride of 2, and padding of 1.

##### Bottleneck

Previous studies [26], [27], [28] have shown the effectiveness of integrating dense connections and deeper networks in SR tasks. To enhance deep feature extraction and fusion, we use cascaded RRDB blocks within a high-dimensional space. This method facilitates the integration of high-dimensional features both before and after deepening the network with dense connections. Meanwhile, residual blocks can help reduce computational overhead and address gradient vanishing issues commonly associated with deeper networks. The proposed method is designed to promote the optimization of critical visual features and facilitate the extraction of detailed features, particularly lines and curves.

##### Decoder

A decoder with progressive fusion is used to merge the encoder and bottleneck features for HR MR image generation. This decoder utilizes convolutional layers with a kernel size of 3 and padding of 1 for upsampling, along with an efficient sub-pixel convolution operation (pixshuffle) at each layer. Skip connections are used to retain the large-scale low-dimensional information obtained from the encoder and perform multi-scale fusion with the small-scale high-dimensional information generated by the bottleneck, thereby increasing the amount of information obtained by the encoder. This allows the model to simultaneously focus on both the large-scale contour structures and small-scale texture details, resulting in more detailed and accurate MR image reconstruction.

### Discriminator

Inspired by [29], we adopt an attention U-net discriminator architecture to specifically target and improve poorly generated textures during adversarial training. This architecture includes attention blocks (AB) and concentration blocks (CB), building upon a U-Net structure, as shown in Fig. 2(e). To boost training stability and tailor the discriminator more effectively for image SR tasks, we apply a spectral normalization regularization operation as described in [30].

### SRN loss

The SRN loss comprises the generator loss and the discriminator loss. We denote the generator and discriminator as *G*(·) and *D*(·), respectively. Specifically, to preserve the structural integrity of MR images and enhance their texture details, we introduce the multi-layer perceptual loss ℒ_*mper*_ into the generator. The pre-trained VGG19 is used to extract image features for the multi-layer perceptual loss, with layers *l* ∈ {2, 7, 9, 12, 17} selected and weighted at *τ*_*l*_ = {0.1, 0.1, 1.0, 1.0, 1.0}. We further employ the 1-norm distance loss ℒ_1_ into the generator to evaluate the distance between the generated HR image and the ground truth. The multi-layer perceptual loss and content distance loss are as follows:

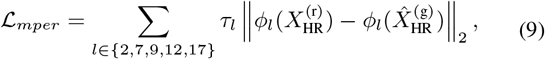

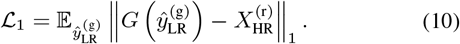

Therefore, the discriminator loss, enhanced generator loss, and the total SRN loss are calculated as follows:

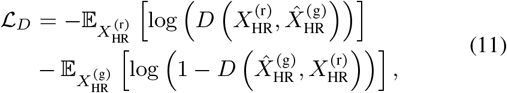

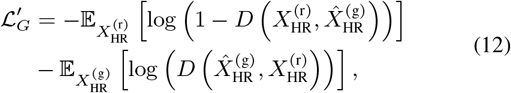

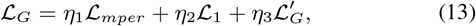

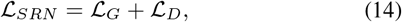

where *η*_1_, *η*_2_ and *η*_3_ are hyperparameters for adjusting different loss terms. We set *η*_1_ = 1, *η*_2_ = 1, and *η*_3_ = 0.1 across the experiments.

## III. Experiments

We utilized two unpaired HR/LR MR image datasets to assess the performance of our proposed method. Specifically, a private HR dataset (i.e., 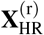) from our institute served as the reference to guide the super-resolution of the LR images 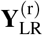, which were sourced from the publicly available Alzheimer’s Disease Neuroimaging Initiative (ADNI) cohort. In addition, we collected paired LR images corresponding to our private HR scans to serve as ground truth for validating the down-sampling ability of the proposed DSN module. Further details on these datasets are provided below.

### A. Experimental setup

#### ADNI LR image dataset

We include a total of 41 T1-weighted 1.5T MRI scans from ADNI Phase 1 (ADNI1) for super-resolution. Although the original resolution of these images, 256×256×160, is not low compared to clinical MR scans, this study aims to achieve a higher sampling density to demonstrate the efficacy of the proposed SR framework. In addition, ADNI1 data is generally considered to have lower image quality compared to the later ADNI phases (i.e., ADNIGO/2 and ADNI 3). Therefore, it motivates us to use ADNI1 images as real LR data and apply our proposed methods to them for both image denoising and SR tasks. From these scans, we extract 3, 520 sagittal images from 22 subjects used for training (as real LR), and 3, 040 images from 19 subjects for testing. Detailed data acquisition protocols can be accessed at adni.loni.usc.edu.

#### Private HR/LR paired image dataset

Four volunteers underwent both HR and LR MRI scanning on a 3.0T GE scanner at our site using a 3D T1-w MP-RAGE sequence. These collected real HR images serve as source references for ADNI LR SR, while our private LR images are used exclusively for DSN validation. The HR scans feature a slice thickness/gap of 1/0 mm, spatial resolution of 1.0 × 1.0 × 1.0 mm, TE of 3.444 ms, TR of 2737.62 ms, and a scanning time of 6 min 48s. The same sequence is used for the LR scanning with a reduced TE of 1.66 ms and scanning time of 4 min 28s. The resolutions of the private HR and LR MRI are 512 × 512 × 388 and 256 × 256 × 388, respectively. As HR and LR MRI share the same field of view (FOV), HR is defined by higher sampling density, whereas LR by lower sampling density. The private HR dataset is divided into the training and testing sets with a ratio of 8 : 2, resulting in 1, 241 images for training and 311 images for testing.

#### Evaluation metrics

In the absence of ground truth (e.g., real ADNI HR images are not available), natural image quality evaluator (NIQE) [31] and blind/referenceless image spatial quality evaluator (BRISQUE) [32] are employed to assess the performance of our proposed DDASR framework. Here, NIQE evaluates image quality by analyzing the structure and content to simulate the perceptual quality of the human visual system. BRISQUE quantifies image quality by analyzing statistical features within the spatial domain of the image.

When ground truth is present (e.g., the private LR images are available for comparing the down-sampling performance), we use the peak signal-to-noise ratio (PSNR) [33] and the structural similarity index (SSIM) [34] to evaluate the performance of our proposed DSN (Sec.III-C). Specifically, PSNR quantifies image quality by comparing the signal-to-noise ratio between the real and synthetic images, while SSIM evaluates similarity from three key aspects: luminance, contrast, and structure.

#### Comparison with prior work

We compare the proposed SR framework with a benchmark method Bicubic, and three state-of-the-art methods including ESRGAN [14], RealSR [35], and RealESRGAN [5]. We further compare our proposed DSN framework with the state-of-the-art DS approaches utilized in ESRGAN, RealSR, and RealESRGAN. Specifically, ESRGAN employs the k-space DS method, RealSR utilizes a degradation framework to estimate blur kernels, and RealESRGAN adopts a high-order degradation modeling method.

To ensure a fair comparison, all compared methods were retrained on our dataset using the same model architectures and optimal hyperparameters as described in their papers. The mean and standard deviation computed across all testing samples are reported for each evaluation metric.

#### Implementation details

Our proposed model is trained on 2 NVIDIA GeForce RTX 3090 GPUs with 48 GB memory using the PyTorch framework. The DSN is trained for 300,000 iterations with a batch size of 3 and a hidden dimension of 512. For the SRN, a cosine annealing with warm restarts learning rate optimization strategy is employed with a minimum learning rate of 1e-7, and the total number of iterations is set to 250,000.

### B. Comparison with the state-of-the-arts

We apply the proposed SR framework on the real ADNI LR data, acknowledged to have lower image quality. Non-reference metrics (NIQE and BRISQUE) are used to evaluate the performance. As illustrated in Table I, our method significantly outperforms others in terms of the NIQE score and achieves the second-best BRISQUE index score. Among the compared SR methods, Bicubic upsampling yields the poorest BRISQUE scores, likely because it simply interpolates the image without considering quality improvement. ESRGAN shows the worst NIQE scores, potentially because it does not take the domain gap into consideration.

**TABLE I.** Performance comparison of ADNI LR images SR.

| Methods | NIQE<sub>std</sub> ↓ | BRISQUE<sub>std</sub> ↓ |
| --- | --- | --- |
| Real LR | 5.18±0.89 | 68.56±6.22 |
| Bicubic | <u>4.91±0.71</u> | 61.31±3.91 |
| ESRGAN | 6.00±0.24 | 60.87±3.92 |
| RealSR | 5.34±0.16 | 57.03±2.13 |
| RealESRGAN | 5.01±0.16 | **40.43±9.43** |
| DDASR (ours) | **4.25±0.15\*** | <u>44.67±3.21</u> |

Best results are in **bold**. The second-best results are underlined. Results with ‘\*’ indicate that our model is significantly better ( $p < 0.05$ ) than the suboptimal method when using the independent t-test.

We further perform a visual comparison of several SR results. As shown in Fig. 3, our proposed method effectively enhances MR image quality by simultaneously preserving structural integrity and recovering biologically significant texture information. RealSR delivers good visual outcomes in certain regions but introduces serious structural alterations and unnecessary texture in many other regions. Although RealESRGAN achieves the best BRISQUE scores, it significantly compromises texture details and suffers from artifacts. Notably, it expands the cortical surface thickness, reduces the depth of brain sulci, and results in unclear boundaries between the cortex and medulla, which may hinder the detection of small lesions.

**Fig. 3.**
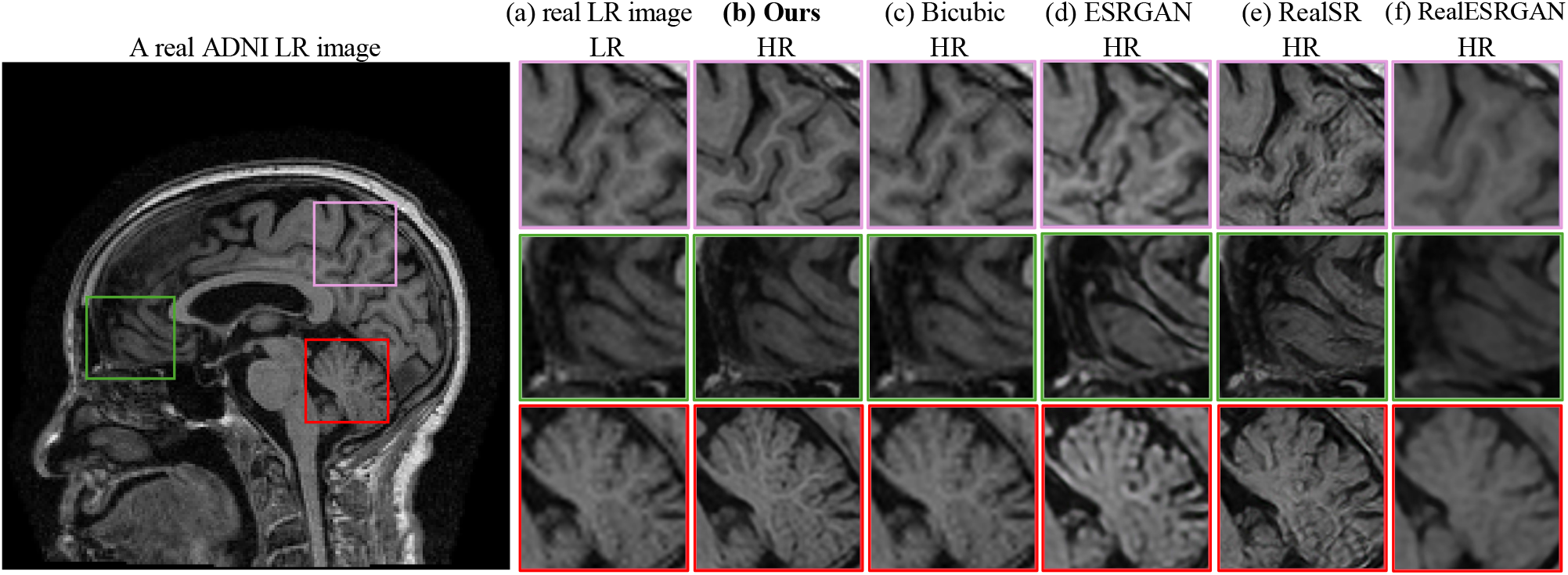
Visual comparison of the SR results from different methods.

### C. Ablation study

We conducted a series of ablation studies to evaluate the effectiveness of the proposed SR framework, the DSN and SRN modules, as well as the key components designed for the SRN. Below, we describe the details of each ablation design and present the corresponding results.

#### DSN: more realistic synthesized LR image

We compare our proposed DSN framework with the DS approaches utilized in RealSR and RealESRGAN, as well as with the k-space method employed by ESRGAN. PSNR and SSIM are used to quantitatively measure the differences between the synthesized LR images and the ground truth (i.e., the real LR images from our site).

As shown in Table II and Fig. 4, our proposed DSN achieves the best performance in terms of PSNR and real-synthetic image distance (i.e., heatmaps in the bottom panel of Fig. 4) and yields the second-best SSIM score. Although k-space obtains the highest SSIM, it actually generates many artifact texture details (see the top panel of Fig. 4(c) and corresponding heatmap). Our DSN outperforms both RealSR and RealESRGAN across all metrics. RealESRGAN produces the poorest PSNR and SSIM scores, possibly due to its down-sampling strategy being optimized for natural images.

**TABLE II.** Performance comparison of the down-sampling methods on the real private LR images.

| Methods | PSNR<sub>std</sub> ↑ | SSIM<sub>std</sub> ↑ |
| --- | --- | --- |
| K-space | 20.52±4.57 | **0.71±0.10** |
| RealESRGAN | 19.98±7.53 | 0.46±0.21 |
| RealSR | <u>20.79±4.37</u> | 0.64±0.08 |
| **Ours** | **21.65±4.21\*** | <u>0.68±0.09</u> |

Best results are in **bold**. The second-best results are underlined. Results with ‘\*’ indicate that our DSN is significantly better ( $P < 0.05$ ) than the suboptimal method when using the independent t-test.

**Fig. 4.**
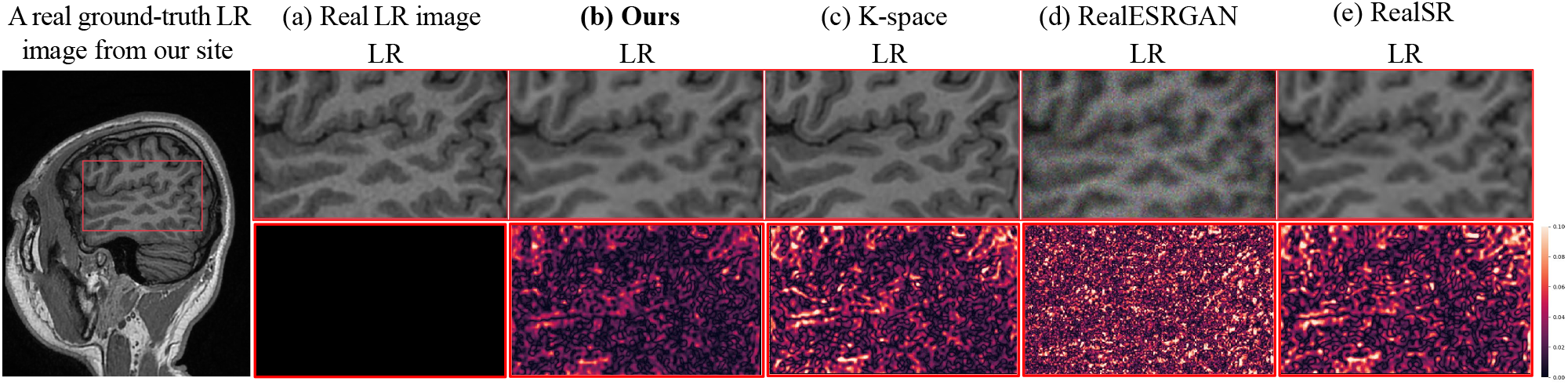
Comparison of the down-sampling methods. A lower heatmap value indicates a smaller distance between the synthesized LR image and the real LR image.

#### Super-resolution network evaluation

We feed the down-sampled LR images generated by each of the benchmark methods into our SRN to examine whether our proposed SR method surpasses the performance of the others. The evaluation is conducted on the ADNI data, employing NIQE and BRISQUE metrics for performance comparison. As shown in Table III, our SRN demonstrates superior performance than the other SR methods regardless of the synthetic LR inputs. This demonstrates the generalizability and stability of our proposed SRN.

**TABLE III.** Performance evaluation of the proposed SRN on ADNI LR IMAGE SR.

| Methods | NIQE<sub>std</sub> ↓ | BRISQUE<sub>std</sub> ↓ |
| --- | --- | --- |
| Bicubic | 4.91±0.71 | 61.31±3.91 |
| Bicubic<sup>#</sup> | 4.33±0.14 | 52.12±3.83 |
| ESRGAN | 6.00±0.24 | 60.87±3.92 |
| ESRGAN<sup>#</sup> | 5.08±0.55 | 103.23±4.30 |
| RealSR | 5.34±0.16 | 57.03±2.13 |
| RealSR<sup>#</sup> | 4.75±0.16 | 54.87±4.32 |
| RealESRGAN | 5.01±0.16 | 40.46±9.43 |
| RealESRGAN<sup>#</sup> | 4.51±0.14 | 35.35±2.83 |

All method utilize their specific downsampling strategy. Methods marked with superscript <sup>#</sup> employ our proposed SRN module, while those without the superscript use their own SRN modules.

#### Evaluation of GAN key components

We further evaluated the effectiveness of the key components designed for the GAN. We accordingly remove the multi-layer perceptual loss (ℒ_*mper*_) from the generator and replace the attention U-Net architecture with the relativistic discriminator (D(RA)) [36].

Ablation experiments are conducted to super-resolve ADNI LR images, utilizing NIQE and BRISQUE metrics for performance comparison. The results, presented in Table IV, indicate that removing the multi-layer perceptual loss leads to a 17.9% reduction in NIQE and a 15.4% reduction in BRISQUE. We also observe that the attention-based U-Net discriminator demonstrates significant advantages, achieving the best results across both evaluation metrics (13.2% BRISQUE and 14.5% NIQE reductions).

**TABLE IV.** Ablation study of the multi-perceptual loss and the attention U-Net mechanism on ADNI LR image SR.

| Methods | NIQE<sub>std</sub> ↓ | BRISQUE<sub>std</sub> ↓ |
| --- | --- | --- |
| w/o $\mathcal{L}_{mper}$ | 5.01±0.21 | 51.54±3.22 |
| w/ $\mathcal{L}_{mper}$ | **4.25±0.15\*** | **44.67±3.21\*** |
| D(RA) | 4.87±0.23 | 50.58±4.44 |
| D(Attention U-Net) | **4.25±0.15\*** | **44.67±3.21\*** |

Best results are in **bold**. Results with ‘\*’ indicate that the key component shows significant improvement (t-test $p < 0.05$ ).

## IV. Conclusion

We propose a novel domain-distance adapted super-resolution framework for MR brain images SR. Our approach combines a transformer-based domain adaptation network, an encoding-decoding generator architecture, an attention UNet-based discriminator for SR, and multi-layer perceptual loss to preserve brain structural and texture information effectively. Experimental results demonstrate the superiority of our proposed model compared to state-of-the-art SR methods in both quantitative evaluation and perceptual quality assessments. This indicates that our proposed framework is a reliable solution for restoring outdated MRI images and has the potential to enhance clinical MR images. The proposed approach has wide applicability to various medical imaging tasks and can assist medical professionals in obtaining more accurate and detailed diagnostic information from 1.5T MR brain images. Therefore, our work has noteworthy contributions to the field of medical imaging and could have a positive impact on patient care.

## Data Availability

All data produced in the present work are contained in the manuscript

https://adni.loni.usc.edu

## ACKNOWLEDGMENT

This work is supported partly by the National Natural Science Foundation of China (62103116, 62102115), the Natural Science Foundation of Heilongjiang Province (LH2022F016), the Fundamental Research Funds for the Central Universities (3072024GH2604), the Shandong Provincial Natural Science Foundation (2022HWYQ-093), Heilongjiang Province basic research business fees for provincial higher education institutions (2023-KYYWF-0217), Harbin Medical University Cancer Hospital Climbing program (PDYS2024-10).

For private HR data, informed consent has been obtained from all participants and the research ethics committees or institutional review boards approved the data collection. ADNI data collection and sharing for this project was funded by the Alzheimer’s Disease Neuroimaging Initiative (ADNI) (National Institutes of Health Grant U01 AG024904) and DOD ADNI (Department of Defense award number W81XWH-12-2-0012).

